# Incidence of outbreak-associated COVID-19 cases by industry in Ontario, Canada, April 1, 2020-March 31, 2021

**DOI:** 10.1101/2021.06.30.21259770

**Authors:** Sarah A. Buchan, Peter M. Smith, Christine Warren, Michelle Murti, Cameron Mustard, JinHee Kim, Sandya Menon, Kevin A. Brown, Trevor van Ingen, Brendan T. Smith

**Author notes:** **Corresponding author**: Sarah Buchan, 661 University Avenue, Floor 17, Toronto, ON, M5G 1M1, Canada.

## Abstract

**Objectives:** The objective of our study was to estimate the rate of workplace outbreak-associated cases of COVID-19 by industry in labour market participants aged 15-69 years who reported working the majority of hours outside the home in Ontario, Canada.

**Methods:** We conducted a population based cross-sectional study of COVID-19 workplace outbreaks and associated-cases reported in Ontario between April 1, 2020 and March 31, 2021. All outbreaks were manually classified into two digit North American Industry Classification System (NAICS) codes. We obtained denominator data from the Statistics Canada Labour Force Survey in order to estimate the incidence of outbreak-associated cases per 100,000,000 hours amongst individuals who reported the majority of hours were worked outside the home. We performed this analysis across industries and in three distinct time periods.

**Results:** Overall, 12% of cases were attributed to workplace outbreaks among working age adults across our study period. While incidence varied across the time periods, the five industries with the highest incidence rates across our study period were agriculture; healthcare and social assistance; food manufacturing; educational services; and, transportation and warehousing.

**Conclusions:** Certain industries have consistently increased incidence of COVID-19 over the course of the pandemic. These results may assist in ongoing efforts to reduce transmission of COVID-19, by prioritizing resources, as well as industry-specific guidance, vaccination, and public health messaging.

## Introduction

Understanding the role of workplace exposure to COVID-19, and differential risk by industry, is critical to reducing morbidity and mortality. Occupational risk is an important source of COVID-19 exposure and transmission[1]. Elevated risk of COVID-19 has been documented among healthcare workers[2], given direct contact with COVID-19 patients[3]. However, workplace outbreaks of COVID-19 have consistently been observed across many industries beyond healthcare, especially in essential services where work is unable to be done from home[4]. A comprehensive analysis of the distribution of workplace outbreaks across industries is important to understand the effectiveness and the limitations of workplace infection prevention and control practices, as well to ensure equitable public health measures (including vaccination) to reduce risk in workplaces and prevent ongoing spread in the community.

The location and frequency of workplace outbreaks will vary by region, depending on the prevalence of industries and intensity of COVID-19 [5]. A number of occupational characteristics have been observed to increase COVID-19 risk at work, including physical proximity to others[6], exposure to disease[7], and indoor ventilation; further, protections in the workplace may vary by industry (Smith et al. forthcoming[8]). In Ontario, an analysis of workplace outbreaks early in the pandemic (between January 21, 2020 to June 30, 2020) found that 68% of outbreaks and 80% of cases belonged to manufacturing, agriculture and transportation warehousing after excluding hospital, congregate living and education and childcare settings[9]. Since this period, Ontario has experienced two additional waves of COVID-19, accompanied by adjustments to public health measures that restricted operations in some industries. As such, it is critical to use accurate denominator data in order to estimate risk of COVID-19 through work. Surveillance systems are often limited in their capture of occupational data [10][11]; however, outbreak data present an opportunity to explore cases associated with reported outbreaks within workplaces in order to mitigate this limitation.

Understanding differences in COVID-19 incidence among workers in industries is required to understand risk and inform prevention practices. The objective of our study was to estimate the rate of workplace outbreak-associated cases of COVID-19 by industry in labour market participants aged 15-69 years who reported working the majority of hours outside the home in Ontario, Canada. We also aimed to estimate the proportion of cases in this age group that were associated with a workplace-associated outbreak.

## Methods

We conducted a population based cross-sectional study of COVID-19 workplace outbreaks and associated-cases reported in Ontario between April 1, 2020 and March 31, 2021. All outbreaks and cases in Ontario are entered into the Public Health Case and Contact Management Solution (CCM), the provincial reportable disease surveillance system, by one of 34 local public health units (PHU). We used monthly data from Ontario respondents to Statistics Canada’s Labour Force Survey (LFS) to estimate the size of the Ontario workforce to quantify the population at risk from April 2020 through March 2021[12]. The LFS is a monthly household survey that uses a rotating panel sample design consisting of six representative panels, where one panel is replaced each month allowing for efficient estimation of monthly changes in the Canadian labour force. LFS respondents are representative of 98% of Canadians aged 15 years and over, excluding persons living on reserves, full-time members of the Canadian Armed Forces, and institutionalized populations[12]. In response to COVID-19 pandemic, a LFS Supplement was introduced in April 2020, which asked questions on where the respondent had spent the majority of their work hours in the previous week (e.g. at home, at the worksite, or outside of the home, but not in a particular location). Questions in the LFS supplement are only asked of respondents aged 15 to 69 years, so we further restricted our sample to COVID-19 cases aged 15-69 years to focus on labour market participants. The Public Health Ontario Ethics Review Board determined that this project did not require research ethics committee approval as the activities described were considered public health practice and not research.

### Outbreak definition and Industry Assignment

In Ontario, PHUs are responsible for declaring COVID-19 outbreaks based on provincial guidance regarding the assessment of risk of acquisition and transmission in a workplace. The outbreak definition varied by industry setting[13], with individual cases constituting an outbreak in long-term care homes (and childcare settings until November 9, 2020), or two cases occurring within 14 days with an epidemiologic link in other settings[14]. For hospitals, long-term care homes, and education settings, outbreaks were classified on PHU entry using existing look up tables available in CCM. All other outbreaks were reviewed retrospectively based on locations (address and outbreak name as entered by the PHU) to ensure consistency with data entry across PHUs and to assign two digit (i.e., sector) North American Industry Classification System (NAICS) industry codes based on a manual lookup[15]. Based on reported outbreaks, 13 categories were examined in our study: agriculture, forestry, fishing and hunting; mining and utilities; construction; manufacturing – food; manufacturing – other; wholesale trade; retail trade; transportation and warehousing; educational services; health care and social assistance; accommodation and food services; public administration; and, other service industries. Additional details on the NAICS and classification of industries are available in **Supplementary Appendix 1**.

### Workplace outbreak-associated cases

We restricted our primary sample to only include workplace outbreak-associated cases. All laboratory-confirmed (i.e., those meeting provincial case definition[16]) COVID-19 cases and hospitalizations were obtained from CCM. For healthcare and congregate care/living settings, we included outbreak-associated cases in workers indicated by an occupational flag in CCM in order to exclude patients or residents. For the education industry, we included all non-students aged over 18 years or had an educational staff flag who were linked to a childcare, elementary or secondary school outbreak. All other cases among the working aged population, defined as ‘non-workplace outbreak-associated cases’ were retained as a comparison group, but were not included in the primary analyses. This group included cases in the community, as well as outbreak-related cases in residents of congregate care/living, and outbreak-related cases in settings where working status data were not available and transmission was unlikely to be restricted to workers only – these included recreational fitness settings (e.g., gyms), other recreational settings (e.g., visual arts class) and places of worship.

### Covariates

We distinguished dates of cases and outbreaks across three time periods: April 1-August 31, 2020 (period 1), September 1 – December 31, 2020 (period 2), and January 1 – March 31, 2021 (period 3). These time periods coincided with changes to public health measures (i.e., stay at home order)[17], the rise of prevalence of variants of concern [18], and allowed for adequate sample size to be obtained from the LFS based on the survey’s sampling strategy [12]. Demographic information on outbreak-associated cases included: gender, age (10-year categories), and diagnosing PHU. Further, quintiles of neighbourhood material deprivation and diversity (measured using the ethnic concentration dimension) were measured using the Ontario Marginalization Index (ON-MARG)[19].

### Statistical Analyses

We examined COVID-19-related cases and hospitalizations across characteristics of workplace and to non-workplace associated cases. Further, we aggregated these outcomes by industry across three time periods. For each period, we estimated industry-specific incidence rates per 100,000,000 work hours and per 100,000 workers who reported the majority of hours were worked outside the home. Respondents were asked ‘the location where the respondent worked the most hours in the previous week’ and our analyses were restricted to individuals reporting the majority of hours were worked outside the home.

We calculated a standardized incidence ratio (SIR) and 95% confidence intervals[20], as the ratio of the workplace outbreak-associated COVID-19 incidence rate to the overall incidence rate in Ontarians aged 15-69, for each industry and time period. We estimated the overall rate by summing the number of COVID-19 cases in Ontario and dividing it by the sum of waking hours (assuming 16 hours of awake time per person per day multiplied by the Ontario population aged 15-69 estimated from projection data for 2020 sourced from Ministry, IntelliHEALTH Ontario).

We performed sensitivity analyses to: 1) include an estimate of temporary foreign workers in agricultural settings who are captured in the case data but not in the LFS denominator[21]; and, 2) reclassify the hours of those self-employed (with employees) on farms to working outside the home (i.e., to ensure their exposure to others was enumerated).

All analyses were conducted in R-Studio (Version 1.2.5019).

## Results

Between April 1, 2020 and March 31, 2021, there were 282,539 COVID-19 cases reported in Ontarians aged 15-69 years. Of these, 247,371 were excluded as they were non-workplace outbreak-associated cases (i.e., cases not associated with an outbreak, residents of congregate care/living, or not meeting workplace associated outbreak definition, **Supplementary Appendix 2 and 3**). Our final study population included 35,168 cases associated with 5,759 workplace outbreaks.

The number of COVID-19 cases and hospitalizations across sociodemographic characteristics by workplace outbreak and non-workplace outbreak-associated cases are presented in **Table 1**. Overall, 12% of cases and 7% of hospitalizations were attributed to workplace outbreaks among working age adults, with 2% and 3% workplace and non-workplace outbreaks-associated cases requiring hospitalization, respectively. Despite an increase in COVID-19 cases and hospitalizations occurring in periods 2 and 3 (September 1 – December 31, 2020 and January 1 – March 31, 2021) compared to period 1 (April 1-August 30, 2020) overall, a lower percentage of workplace compared to non-workplace outbreak-associated cases and hospitalization were observed. The proportion of workplace outbreak-associated cases was higher among females (14%) compared to males (11%) but hospitalizations were similar across gender. The proportion of workplace outbreak-associated cases differed by geography (i.e. PHU), ranging from approximately 5% of all cases among the working population to 27% of all cases, depending on the PHU.

**Table 1:**
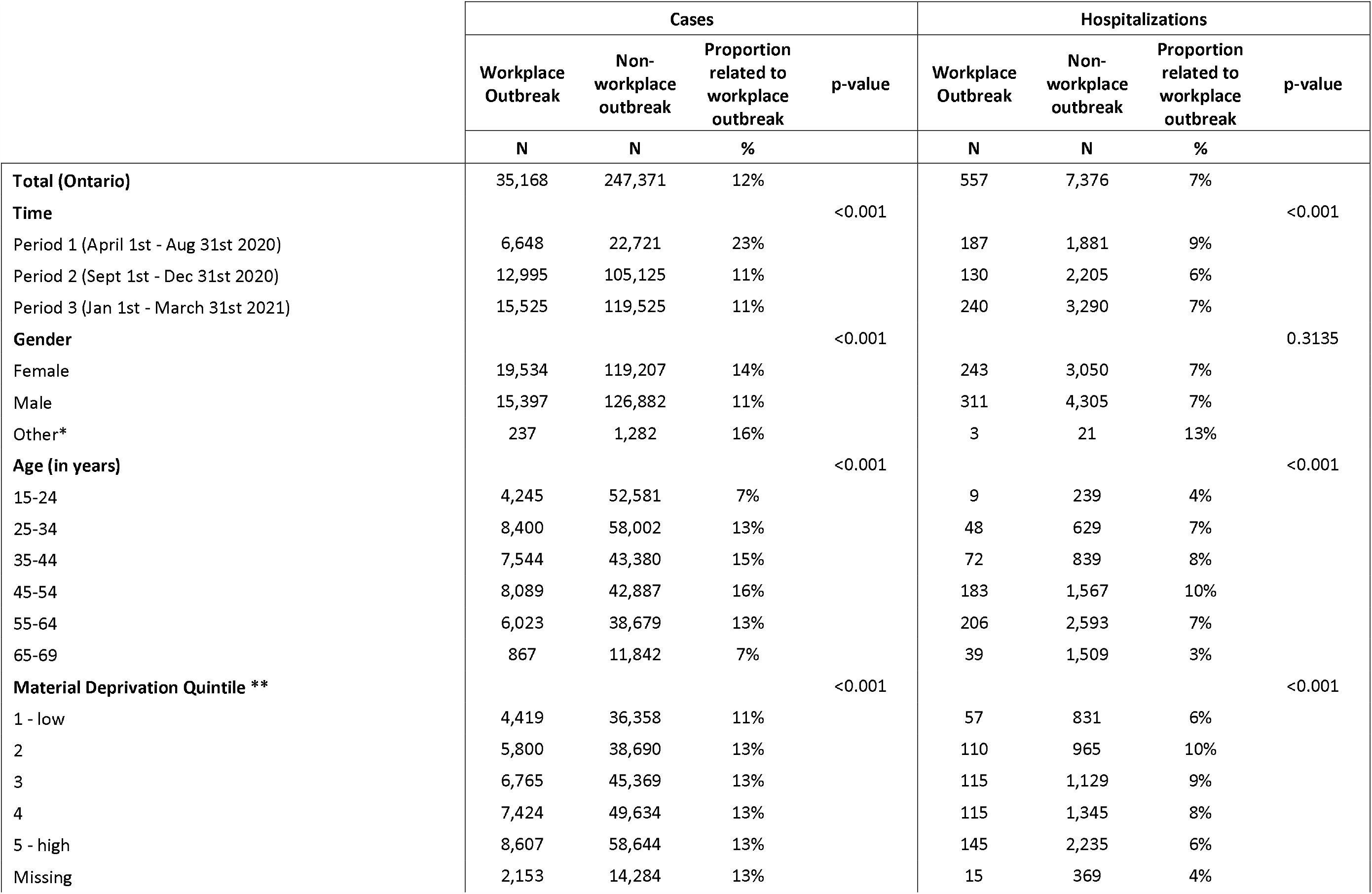
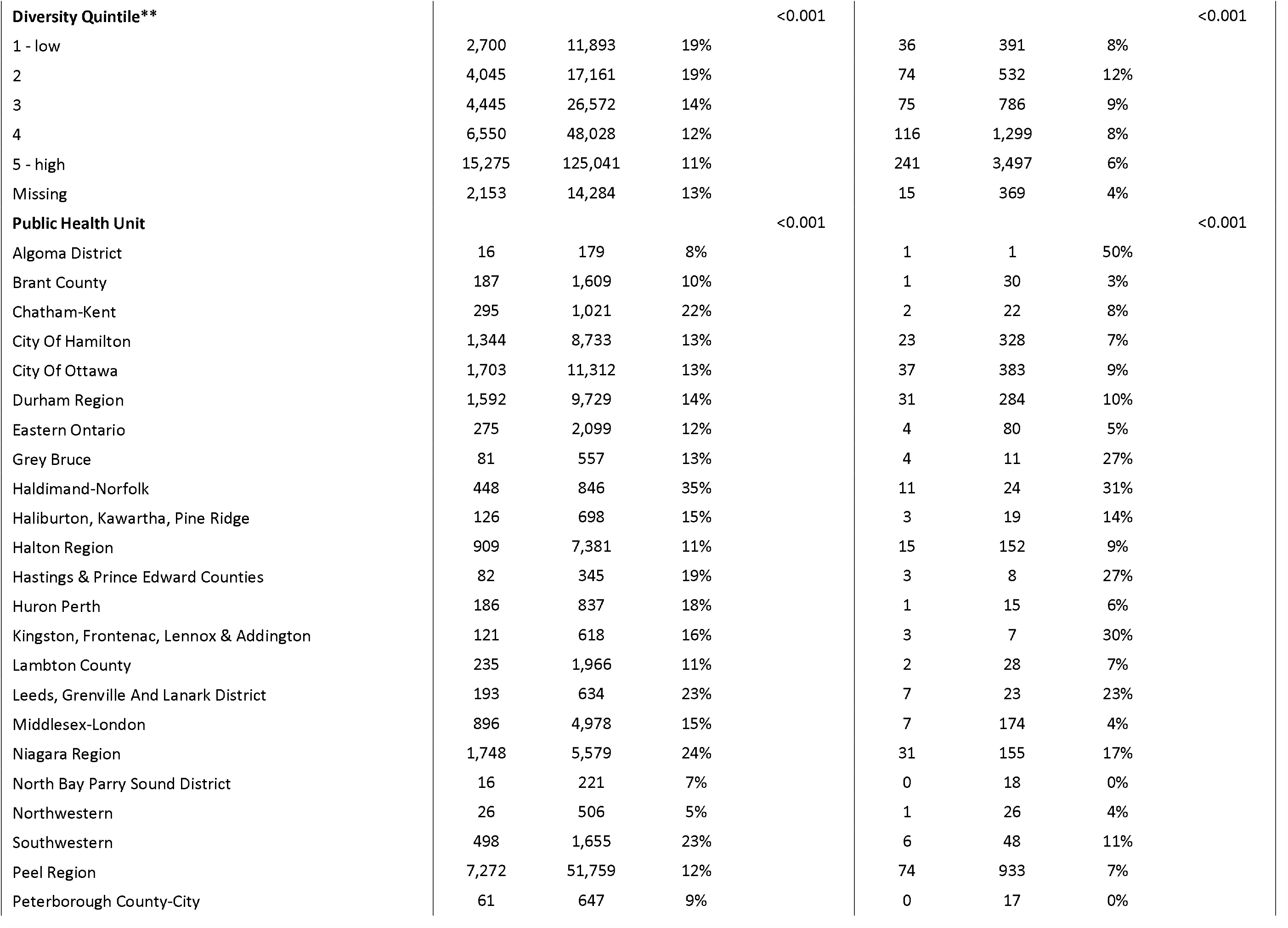
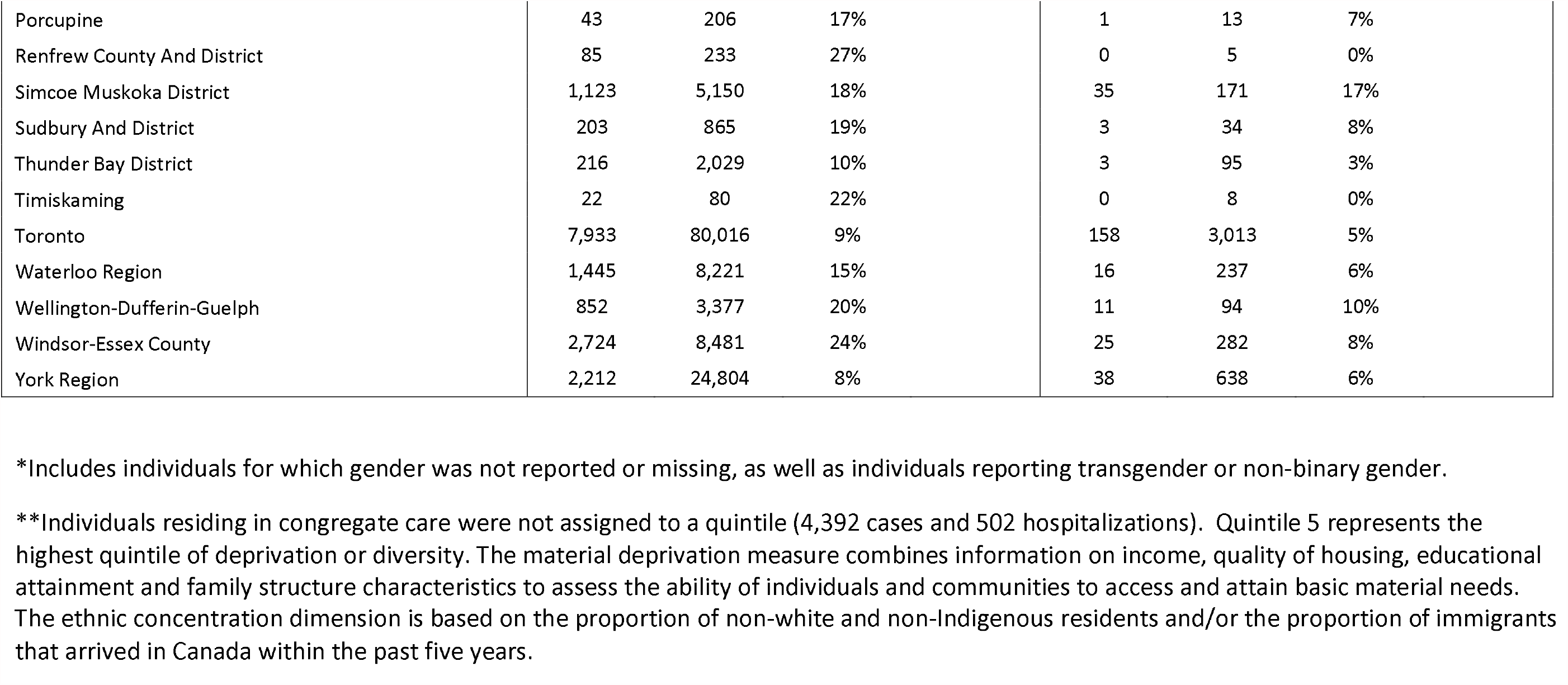
Socio-demographic characteristics of COVID-19 cases and hospitalizations among 15-69 year olds, reported April 1, 2020 - March 31, 2021 in workplace and non-workplace outbreak-associated cases in Ontario, Canada.

|  | Cases |  |  |  | Hospitalizations |  |  |  |
| --- | --- | --- | --- | --- | --- | --- | --- | --- |
|  | Workplace<br>Outbreak | Non-workplace<br>outbreak | Proportion<br>related to<br>workplace<br>outbreak | p-value | Workplace<br>Outbreak | Non-workplace<br>outbreak | Proportion<br>related to<br>workplace<br>outbreak | p-value |
|  | N | N | % |  | N | N | % |  |
| **Total (Ontario)** | 35,168 | 247,371 | 12% |  | 557 | 7,376 | 7% |  |
| **Time** |  |  |  | <0.001 |  |  |  | <0.001 |
| Period 1 (April 1st - Aug 31st 2020) | 6,648 | 22,721 | 23% |  | 187 | 1,881 | 9% |  |
| Period 2 (Sept 1st - Dec 31st 2020) | 12,995 | 105,125 | 11% |  | 130 | 2,205 | 6% |  |
| Period 3 (Jan 1st - March 31st 2021) | 15,525 | 119,525 | 11% |  | 240 | 3,290 | 7% |  |
| **Gender** |  |  |  | <0.001 |  |  |  | 0.3135 |
| Female | 19,534 | 119,207 | 14% |  | 243 | 3,050 | 7% |  |
| Male | 15,397 | 126,882 | 11% |  | 311 | 4,305 | 7% |  |
| Other* | 237 | 1,282 | 16% |  | 3 | 21 | 13% |  |
| **Age (in years)** |  |  |  | <0.001 |  |  |  | <0.001 |
| 15-24 | 4,245 | 52,581 | 7% |  | 9 | 239 | 4% |  |
| 25-34 | 8,400 | 58,002 | 13% |  | 48 | 629 | 7% |  |
| 35-44 | 7,544 | 43,380 | 15% |  | 72 | 839 | 8% |  |
| 45-54 | 8,089 | 42,887 | 16% |  | 183 | 1,567 | 10% |  |
| 55-64 | 6,023 | 38,679 | 13% |  | 206 | 2,593 | 7% |  |
| 65-69 | 867 | 11,842 | 7% |  | 39 | 1,509 | 3% |  |
| **Material Deprivation Quintile \*\*** |  |  |  | <0.001 |  |  |  | <0.001 |
| 1 - low | 4,419 | 36,358 | 11% |  | 57 | 831 | 6% |  |
| 2 | 5,800 | 38,690 | 13% |  | 110 | 965 | 10% |  |
| 3 | 6,765 | 45,369 | 13% |  | 115 | 1,129 | 9% |  |
| 4 | 7,424 | 49,634 | 13% |  | 115 | 1,345 | 8% |  |
| 5 - high | 8,607 | 58,644 | 13% |  | 145 | 2,235 | 6% |  |
| Missing | 2,153 | 14,284 | 13% |  | 15 | 369 | 4% |  |
| **Diversity Quintile\*\*** |  |  |  | <0.001 |  |  |  | <0.001 |
| 1 - low | 2,700 | 11,893 | 19% |  | 36 | 391 | 8% |  |
| 2 | 4,045 | 17,161 | 19% |  | 74 | 532 | 12% |  |
| 3 | 4,445 | 26,572 | 14% |  | 75 | 786 | 9% |  |
| 4 | 6,550 | 48,028 | 12% |  | 116 | 1,299 | 8% |  |
| 5 - high | 15,275 | 125,041 | 11% |  | 241 | 3,497 | 6% |  |
| Missing | 2,153 | 14,284 | 13% |  | 15 | 369 | 4% |  |
| **Public Health Unit** |  |  |  | <0.001 |  |  |  | <0.001 |
| Algoma District | 16 | 179 | 8% |  | 1 | 1 | 50% |  |
| Brant County | 187 | 1,609 | 10% |  | 1 | 30 | 3% |  |
| Chatham-Kent | 295 | 1,021 | 22% |  | 2 | 22 | 8% |  |
| City Of Hamilton | 1,344 | 8,733 | 13% |  | 23 | 328 | 7% |  |
| City Of Ottawa | 1,703 | 11,312 | 13% |  | 37 | 383 | 9% |  |
| Durham Region | 1,592 | 9,729 | 14% |  | 31 | 284 | 10% |  |
| Eastern Ontario | 275 | 2,099 | 12% |  | 4 | 80 | 5% |  |
| Grey Bruce | 81 | 557 | 13% |  | 4 | 11 | 27% |  |
| Haldimand-Norfolk | 448 | 846 | 35% |  | 11 | 24 | 31% |  |
| Haliburton, Kawartha, Pine Ridge | 126 | 698 | 15% |  | 3 | 19 | 14% |  |
| Halton Region | 909 | 7,381 | 11% |  | 15 | 152 | 9% |  |
| Hastings & Prince Edward Counties | 82 | 345 | 19% |  | 3 | 8 | 27% |  |
| Huron Perth | 186 | 837 | 18% |  | 1 | 15 | 6% |  |
| Kingston, Frontenac, Lennox & Addington | 121 | 618 | 16% |  | 3 | 7 | 30% |  |
| Lambton County | 235 | 1,966 | 11% |  | 2 | 28 | 7% |  |
| Leeds, Grenville And Lanark District | 193 | 634 | 23% |  | 7 | 23 | 23% |  |
| Middlesex-London | 896 | 4,978 | 15% |  | 7 | 174 | 4% |  |
| Niagara Region | 1,748 | 5,579 | 24% |  | 31 | 155 | 17% |  |
| North Bay Parry Sound District | 16 | 221 | 7% |  | 0 | 18 | 0% |  |
| Northwestern | 26 | 506 | 5% |  | 1 | 26 | 4% |  |
| Southwestern | 498 | 1,655 | 23% |  | 6 | 48 | 11% |  |
| Peel Region | 7,272 | 51,759 | 12% |  | 74 | 933 | 7% |  |
| Peterborough County-City | 61 | 647 | 9% |  | 0 | 17 | 0% |  |

|  |  |  |  |  |  |  |
| --- | --- | --- | --- | --- | --- | --- |
| Porcupine | 43 | 206 | 17% | 1 | 13 | 7% |
| Renfrew County And District | 85 | 233 | 27% | 0 | 5 | 0% |
| Simcoe Muskoka District | 1,123 | 5,150 | 18% | 35 | 171 | 17% |
| Sudbury And District | 203 | 865 | 19% | 3 | 34 | 8% |
| Thunder Bay District | 216 | 2,029 | 10% | 3 | 95 | 3% |
| Timiskaming | 22 | 80 | 22% | 0 | 8 | 0% |
| Toronto | 7,933 | 80,016 | 9% | 158 | 3,013 | 5% |
| Waterloo Region | 1,445 | 8,221 | 15% | 16 | 237 | 6% |
| Wellington-Dufferin-Guelph | 852 | 3,377 | 20% | 11 | 94 | 10% |
| Windsor-Essex County | 2,724 | 8,481 | 24% | 25 | 282 | 8% |
| York Region | 2,212 | 24,804 | 8% | 38 | 638 | 6% |

\*Includes individuals for which gender was not reported or missing, as well as individuals reporting transgender or non-binary gender.
\*\*Individuals residing in congregate care were not assigned to a quintile (4,392 cases and 502 hospitalizations). Quintile 5 represents the highest quintile of deprivation or diversity. The material deprivation measure combines information on income, quality of housing, educational attainment and family structure characteristics to assess the ability of individuals and communities to access and attain basic material needs. The ethnic concentration dimension is based on the proportion of non-white and non-Indigenous residents and/or the proportion of immigrants that arrived in Canada within the past five years.

The number of workplace outbreak-associated COVID-19 outbreaks, cases and hospitalizations and SIRs by industry and time period are presented in **Table 2.** A SIR greater than 1.0 indicates that there was a higher rate of COVID-19 cases per hour exposed in a given industry compared to what was observed in the overall working age population, while a SIR less than 1.0 indicates a decreased rate. The majority of workplace-associated cases were attributed to select industries; these industries were consistent over time but the distribution varied between periods and was impacted by public health measures. In period 1, excess workplace outbreak-associated cases (SIR) were observed in agriculture (24.9), healthcare and social assistance (9.3) and food manufacturing (5.0) industries. Similar trends were observed in period 2 and 3, although to a lesser extent, with cases 2.4 and 4.3 times higher in agriculture, 2.6 and 2.2 times higher in health care and social assistance and 2.6 and 2.4 times higher in food manufacturing industries. In addition, excess cases were observed in transportation and warehousing (period 2: 1.1; period 3: 1.5) and education (period 1: 1.2; period 3: 1.1) industries. The incidence of workplace outbreak-associated COVID-19 cases per 100,000,000 hours worked by industry and time period are presented in **Figure 1**.

**Table 2:** COVID-19 cases and hospitalizations workplace outbreak-associated cases and standardized incidence ratio (SIR), by industry and period among workers aged 15-69 in Ontario, Canada reported April 1, 2020 – March 31, 2021.

| Time period & Industry | Workplace outbreaks | Cases |  | Hospitalizations |  | SIR (95%CI) |
| --- | --- | --- | --- | --- | --- | --- |
|  | N | N | % | N | % |  |
| **<u>Period 1 (April 1st - Aug 31st 2020)</u>** |  |  |  |  |  |  |
| Accommodation and Food Service | 16 | 49 | 1% | 4 | 2% | 0.4 (0.3, 0.6) |
| Agriculture | 29 | 1,339 | 20% | 21 | 11% | 24.9 (23.5, 24.9) |
| Construction | 11 | 43 | 1% | 0 | 0% | 0.1 (0.1, 0.4) |
| Education | 17 | 45 | 1% | 0 | 0% | 1.2 (0.9, 1.5) |
| Healthcare and Social Assistance | 549 | 4,050 | 61% | 130 | 70% | 9.3 (9.0, 9.3) |
| Manufacturing - Food | 32 | 474 | 7% | 17 | 9% | 5.0 (4.6, 5.1) |
| Manufacturing - Other | 63 | 313 | 5% | 6 | 3% | 0.8 (0.7, 0.9) |
| Mining and Utilities | 1 | 21 | 0% | 1 | 1% | 0.5 (0.3, 0.9) |
| Other Service Industries | 19 | 70 | 1% | 1 | 1% | 0.1 (0.1, 0.4) |
| Public Administration | 5 | 32 | 0% | 3 | 2% | 0.3 (0.2, 0.6) |
| Retail Trade | 16 | 42 | 1% | 0 | 0% | 0.1 (0.1, 0.4) |
| Transportation and Warehousing | 29 | 164 | 2% | 4 | 2% | 0.8 (0.7, 0.9) |
| Wholesale Trade | 3 | 6 | 0% | 0 | 0% | 0.1 (0.0, 0.9) |
| **<u>Period 2 (Sept 1st - Dec 31st 2020)</u>** |  |  |  |  |  |  |
| Accommodation and Food Service | 114 | 528 | 4% | 6 | 5% | 0.6 (0.6, 0.7) |
| Agriculture | 26 | 532 | 4% | 4 | 3% | 2.4 (2.2, 2.5) |
| Construction | 58 | 192 | 1% | 2 | 2% | 0.1 (0.1, 0.3) |
| Education | 445 | 923 | 7% | 8 | 6% | 0.8 (0.8, 0.9) |
| Healthcare and Social Assistance | 1,113 | 5,862 | 45% | 60 | 46% | 2.6 (2.6, 2.7) |
| Manufacturing - Food | 72 | 861 | 7% | 5 | 4% | 2.6 (2.4, 2.7) |
| Manufacturing - Other | 214 | 1,577 | 12% | 15 | 12% | 0.8 (0.7, 0.8) |
| Mining and Utilities | 6 | 19 | 0% | 0 | 0% | 0.1 (0.1, 0.5) |
| Other Service Industries | 100 | 457 | 4% | 7 | 5% | 0.2 (0.2, 0.3) |
| Public Administration | 31 | 130 | 1% | 1 | 1% | 0.3 (0.2, 0.5) |
| Retail Trade | 96 | 528 | 4% | 7 | 5% | 0.3 (0.3, 0.4) |
| Transportation and Warehousing | 64 | 1,153 | 9% | 12 | 9% | 1.1 (1.0, 1.1) |
| Wholesale Trade | 32 | 233 | 2% | 3 | 2% | 0.5 (0.4, 0.6) |
| **Period 3 (Jan 1st - March 31st 2021)** |  |  |  |  |  |  |
| Accommodation and Food Service | 88 | 391 | 3% | 9 | 4% | 0.5 (0.5, 0.6) |
| Agriculture | 78 | 705 | 5% | 9 | 4% | 4.3 (4.0, 4.4) |
| Construction | 118 | 562 | 4% | 6 | 3% | 0.3 (0.3, 0.4) |
| Education | 484 | 1,138 | 7% | 20 | 8% | 1.1 (1.1, 1.2) |
| Healthcare and Social Assistance | 1,011 | 5,471 | 35% | 51 | 21% | 2.2 (2.2, 2.3) |
| Manufacturing - Food | 65 | 991 | 6% | 27 | 11% | 2.4 (2.2, 2.4) |
| Manufacturing - Other | 267 | 2,450 | 16% | 56 | 23% | 1.1 (1.0, 1.1) |
| Mining and Utilities | 14 | 75 | 0% | 1 | 0% | 0.3 (0.2, 0.5) |
| Other Service Industries | 146 | 688 | 4% | 19 | 8% | 0.3 (0.3, 0.4) |
| Public Administration | 53 | 376 | 2% | 5 | 2% | 0.7 (0.6, 0.8) |
| Retail Trade | 128 | 718 | 5% | 11 | 5% | 0.3 (0.3, 0.4) |
| Transportation and Warehousing | 106 | 1,739 | 11% | 22 | 9% | 1.5 (1.4, 1.5) |
| Wholesale Trade | 39 | 221 | 1% | 4 | 2% | 0.4 (0.3, 0.5) |

**Figure 1.**
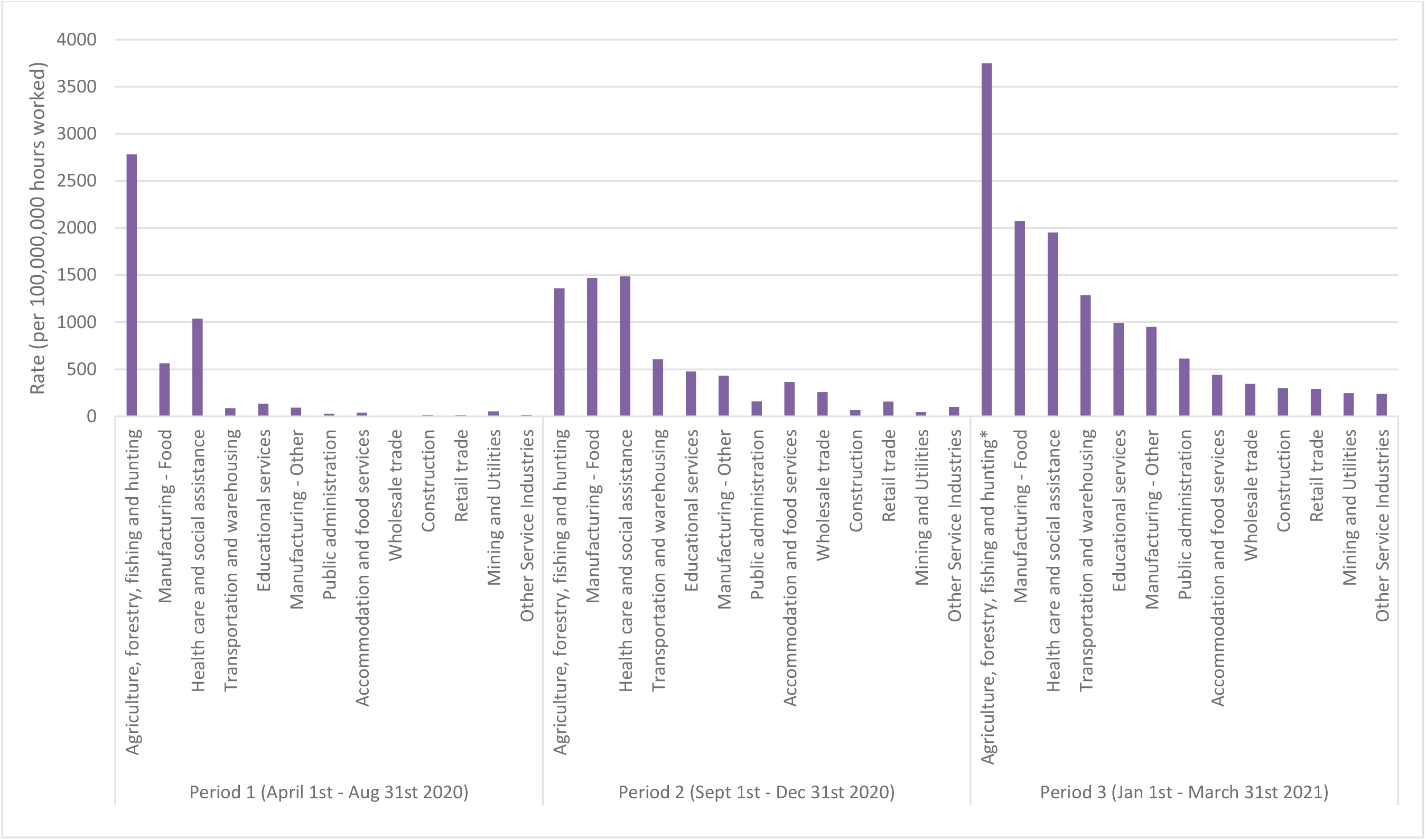
Cumulative case rate (per 100,000,000 hours worked outside the home) of COVID-19 among Ontario workers aged 15-69 by industry and period.

The incidence of workplace outbreak-associated COVID-19 cases per 100,000 workers by industry and time period are presented in **Supplementary Appendix 4**. The distribution of COVID-19 incidence rates were consistent across industries using both the number of workers and hours worked as denominators.

### Sensitivity analyses

When we updated our results to account for the seasonal variation of temporary foreign workers in agricultural settings and to account for the home also being the work setting for self-employed agriculture workers, the incidence in the agricultural setting decreased in all time periods (**Supplementary Appendix 5)**. However, the ranking of incidence as compared to other industries did not change.

## Discussion

In a population based study including all workplace outbreaks and their associated cases in Ontario, Canada between April 2020 and March 2021, we observed that workplace outbreak-associated cases accounted for 12% of all cases in the working age population. When broken down by industry, incidence rates were highest in healthcare and social assistance, food manufacturing, agriculture, other manufacturing, educational services, and transportation and warehousing. This reflects only cases linked to identified and reported workplace outbreaks and does not account for non-outbreak cases in workers or further spread within households related to index cases associated with workplace outbreaks; as such, the total number of cases resulting from workplace outbreaks is likely to be larger than what is presented in this study[9][22].

Our work is an expansion of previously published estimates for Ontario’s first wave[9], for which denominator data were not available. That study coincides with period 1 in our analysis, reflecting a time when restrictions for essential services were stronger than during subsequent periods and when asymptomatic testing was limited. Similar to these previous results, we found a high incidence of outbreak-related cases in manufacturing (including food), agriculture, and transportation and warehousing industries. In our updated results, we also found high incidence (based on hours worked outside the home) in the education industry during periods that included timeframes when schools had reopened for in-person learning. The incidence across industries was highest in the third period of our study, which encompassed the peak of the second wave and beginning of the third wave of COVID-19 in Ontario[18]. This period also overlapped with the rapid rise of variants of concern (predominantly B.1.1.7), which are not only more transmissible but which also led to the lowering of thresholds for contact and outbreak management, and thereby could have resulted in increased case detection. This period also coincided with the roll out of COVID-19 vaccines to staff working in hospitals and long-term care homes [23], which may explain the comparatively smaller increase in rates for healthcare and social assistance between periods 3 and 2 relative to other industries.

The majority of published estimates report on specific occupations[24][25][26] or industries of interest, particularly healthcare [3] and food processing[27][28]. Other studies have focused on ecological comparisons of rates in neighbourhoods by proportion employed in ‘essential work’[29], but were unable to assess risk across occupations or industries. Few other papers have comprehensively estimated incidence across all industries. Independent analyses of workplace outbreaks by industry in Utah from March to June 2020[30] and Los Angeles County from March to September 2020[31] assessing incidence using NAICS classification and both identified manufacturing, wholesale trade, transportation and warehousing and construction (Utah only) as industries with the highest incidence during these time frames. Chen et al.’s[32] recent analysis in the UK also identified workplace outbreaks by geographical region and industrial sector from May to October 2020 and reported the highest outbreak rate in food manufacturing, followed by warehouses and other manufacturing.

These studies all excluded a combination of healthcare, congregate-living and education settings and included denominator data from 2019 to estimate incidence within their industry classifications, which are unlikely to accurately reflect labour force participation during the pandemic period, given closures to industries and the large proportion of workers currently working remotely (which varies by industry). However, similar to these studies, we identified manufacturing industries as having some of the highest rates of COVID-19, but separated food manufacturing from other manufacturing. Our results demonstrate higher incidence of outbreak-associated COVID-19 in food manufacturing relative to all other manufacturing and align with other studies that have identified outbreaks in food processing facilities [27][28]. Factors that relate to higher risk of COVID-19, including high-density settings, close proximity, prolonged duration of contact, may be particularly prevalent in manufacturing settings[33].

Comparisons to other studies across industries are challenging due to differences in study methodology and data sources (e.g., compensation claims[34], time frames, use of occupational versus industry data [24][25][26], and restrictions in place for the specified geography, including workplace closures. Further, industry, occupation as well as other sociodemographic data on cases and contacts, is often limited in surveillance data. For example, we were unable to disentangle industry-specific risk from the role of other factors in our data, such as occupational risk (including within industries), socioeconomic and racial inequities, household size, and financial barriers to isolate, all of which may be associated with increased risk of COVID-19 [35]. Improved occupational surveillance for COVID-19, along with the collection of other socioeconomic determinants[36], would enhance capabilities to effectively respond to COVID-19 as well as future pandemics[4][5].

## Strengths and Limitations

Our study is not without limitations. We restricted our analyses to workplace outbreak-associated cases; as a result, they do not represent overall rates of COVID-19 among workers. Additionally, not all included outbreak-associated cases were acquired in the workplace, or while on duty – we were unable to distinguish risks incurred in work areas versus work-related activities or circumstances, e.g., carpooling, mealtimes or breaks. There were also likely differences in declaring/managing outbreaks across the study period (e.g., due to capacity for contact tracing and identifying common exposures, access to testing for outbreaks) and by PHU. This would have impacted the overall number of cases linked to workplace outbreaks and their proportion of total cases. Workplace-outbreak related guidance was issued in June 2020 and updated in February 2021 to a lower threshold for identifying contacts for testing and quarantine in the context of variants of concern; as such, there may be additional inconsistency across periods in our study [14]. Further, there may be differential identification of outbreaks across industries. First, enhanced testing initiatives (including certain funded testing programs) [37] implemented in some industries (i.e., healthcare, education) may have led to increased case and outbreak identification. Second, outbreak definitions were not consistent across industries and some changed over time. For example, a single case constituted an outbreak in long-term care settings which may have inflated outbreak-associated cases in the healthcare relative to other industries whereas an outbreak required two epidemiologically linked cases. Another example includes the agricultural industry, for which many outbreaks also had a component of congregate living given many staff reside in provided accommodation. We were unable to distinguish infections acquired in the workplace from those due to co-habitating workers; this factor may be a significant driver of the high incidence in this industry. We have underestimated the incidence in industries where settings were excluded[38], such as gyms and places of worship, where outbreaks were less likely to have been restricted to staff only based on what is known about transmission dynamics in these settings[39][40]. Third, public health measures and interventions changed over the study period and would have impacted the likelihood of transmission in the workplace. For example, in Ontario, schools were closed from March to September 2020 and healthcare workers were among the highest prioritized during the vaccination roll-out (beginning December 2020). Thus, the changing context of closures and other interventions during the periods makes it challenging to compare the estimated incidence across time periods.

Additional limitations include that the data are reflective of an individual’s self-reported main and inability to include data from March 2020 as LFS denominator data were only available as of April 2020. However, few workplace outbreak-associated cases were reported during this month. Finally, there may have been some misclassification related to outbreaks being classified manually into industry; however, as we reported outbreaks at the two digit level, we believe this is minimal.

Our study also has several strengths. First, we were able to estimate the incidence of all workplace outbreak-associated cases, a limitation to previous studies that use general population cohorts (less representative and higher SES [11] or only include information on specific settings. While this approach may not have captured all workplace associated cases, declaration of a workplace outbreak is an indication that workplace transmission was considered reasonable [14]. By using a combination of risk factors in the provincial surveillance system, along with the manual classification of settings and industry, we were able to create a comprehensive dataset of all workplace outbreak-associated cases. This has allowed us to examine industry-specific incidence, including comparisons between non-healthcare and healthcare industries, responding to the stated need to quantify the COVID-19 burden on all workers [4]. These results provide a unique perspective of workplace COVID-19 transmission across all industries, over a time period that spans more than the initial phase of the pandemic for a jurisdiction of over 14 million individuals. Given the severity of restrictions in many jurisdictions during the first half of 2020, results from this time period cannot be extrapolated to other time periods. Second, our analyses incorporate denominator data from 2020/21 and are more reflective of the changes in the number of individuals actually employed, and working outside of the home within an industry during the pandemic than those that rely on older estimates. This stratification mitigates concerns in comparing incidence by restrictions on certain industries, as we have estimated incidence in those individuals who worked outside the home and could therefore be considered ‘at-risk’.

Our results demonstrate that cases associated with workplace-outbreaks continue to contribute to the burden of COVID-19 in working-aged populations in Ontario, although a considerable proportion of COVID-19 cases among working-aged adults in Ontario are not associated with workplace outbreaks. We have also shown that under varying circumstances of changing restrictions and policy guiding outbreak declaration/management, certain industries consistently had increased incidence of COVID-19 over the course of the pandemic. These results may assist in ongoing efforts to reduce transmission of COVID-19, by prioritizing resources, as well as industry-specific guidance, vaccination, and public health messaging.

## Data Availability

Public Health Ontario (PHO) cannot disclose the underlying data. Doing so would compromise individual privacy contrary to PHO's ethical and legal obligations. Restricted access to the data may be available under conditions prescribed by the Ontario Personal Health Information Protection Act, 2004, the Ontario Freedom of Information and Protection of Privacy Act, the Tri-Council Policy Statement: Ethical Conduct for Research Involving Humans (TCPS 2 (2018)), and PHO privacy and ethics policies. Data are available for researchers who meet PHO's criteria for access to confidential data. Information about PHO's data access request process is available on-line at https://www.publichealthontario.ca/en/data-and-analysis/using-data/data-requests.
Access to the anonymized microdata for the Labour Force Survey Supplement is available through Statistics Canada to accredited researchers and government employees for research purposes.

## Acknowledgements

We thank the staff at the Statistics Canada’s Toronto Research Data Centre for their assistance in accessing the data to the LFS and vetting the output for this project.

## Author Contribution

SAB, PMS and BTS designed the study. SAB, SM and TVI extracted surveillance data on cases from CCM and classified outbreaks by industry while PMS extracted denominator data from the Labour Force Survey. CW conducted all the data analysis. SAB, CW and BTS drafted the manuscript. SAB, PMS, CW, MM, CM, JHK, SM, KAB, TVI and BTS all contributed to the interpretation of the data, revising the manuscript and final approval.

## Competing Interest

The authors have declared no competing interest.

## Funding

This study was supported by Public Health Ontario.

The Institute for Work & Health is supported through funding from the Ontario Ministry of Labour, Training and Skills Development (MLTSD). The analyses, conclusions, opinions and statements expressed herein are solely those of the authors and do not reflect those of the MLTSD; no endorsement is intended or should be inferred.

## Data Sharing

Public Health Ontario (PHO) cannot disclose the underlying data. Doing so would compromise individual privacy contrary to PHO’s ethical and legal obligations. Restricted access to the data may be available under conditions prescribed by the Ontario Personal Health Information Protection Act, 2004, the Ontario Freedom of Information and Protection of Privacy Act, the Tri-Council Policy Statement: Ethical Conduct for Research Involving Humans (TCPS 2 (2018)), and PHO privacy and ethics policies. Data are available for researchers who meet PHO’s criteria for access to confidential data. Information about PHO’s data access request process is available on-line at https://www.publichealthontario.ca/en/data-and-analysis/using-data/data-requests.

Access to the anonymized microdata for the Labour Force Survey Supplement is available through Statistics Canada to accredited researchers and government employees for research purposes.

## Ethical Approval

The Public Health Ontario Ethics Review Board determined that this project did not require research ethics committee approval as the activities described were considered public health practice and not research.

