## Supplementary Data for "Incidence of outbreak-associated COVID-19 cases by industry in Ontario, Canada, April 1, 2020-March 31, 2021"

**Supplementary Appendix 1: Supplementary methods for NAICS classification**

The North American Industry Classification System (NAICS)-2017 is a hierarchical classification system developed by Canada, The United States, and Mexico [1]. For our study objective, we classified workplace outbreaks using the 2-digit NAICS industry sectors (N=20), with some modifications (see table below).

First, we separated the NAICS-2017 industry sector 31-33 - manufacturing into 2 distinct groups: food manufacturing (i.e., NAICS-2017 industry subsectors: 311 - food manufacturing and 312 - beverage and tobacco product manufacturing) and other manufacturing. Second, we combined sectors with few outbreaks, including ‘mining and utilities’ (i.e., NAICS-2017 industry sector codes: 21 - mining, quarrying, and oil and gas extraction and 22 – utilities) and ‘other services industries’ (i.e., NAICS-2017 industry sector codes:  51 - information and cultural, 52 - finance and insurance, 53 - real estate, rental and leasing, 54-professional, scientific and technical services, 55-management of companies and enterprises, 56- administrative and support, waste management and remediation services, 71 - arts, entertainment and recreation and 81 - other services (except public administration)). For consistency, we excluded workers from our denominators from industries that were ineligible for our workplace outbreak-associated case definition, including NAICS-2017 industry groups: 6116 - other schools and instruction, 7111 - performing arts companies, 7112 - spectator sports, 7113 - promoters (presenters) of performing arts, sports and similar events, 7114 - agents and managers for artists, athletes, entertainers and other public, 7115 - independent artists, writers and performers, 7131 - amusement parks and arcades 7132 - gambling industries, 7139 - other amusement and recreation industries, and 8131 - religious organizations. Overall, 13 sector categories were examined: agriculture, forestry, fishing and hunting; mining and utilities; construction; manufacturing – food; manufacturing – other; wholesale trade; retail trade; transportation and warehousing; educational services; health care and social assistance; accommodation and food services; public administration; and, other service industries.

**North American Industry Classification System (NAICS) 2017 industry sectors and modifications for study objectives.**

| **North American Industry Classification System (NAICS) 2017: 2-digit Industry Sector** | **Final Industry Sectors in Analysis (N=13)** |
| --- | --- |
| 11 Agriculture, forestry, fishing and hunting | Agriculture, forestry, fishing and hunting |
| 21 Mining, quarrying, and oil and gas extraction | Mining and utilities |
| 22 Utilities | Mining and utilities |
| 23 Construction | Construction |
| 31-33 Manufacturing | Manufacturing - food  Includes: 311 - Food manufacturing and 312 - Beverage and tobacco product manufacturing |
|  | Manufacturing - other |
| 41 Wholesale trade | Wholesale trade |
| 44-45 Retail trade | Retail trade |
| 48-49 Transportation and warehousing | Transportation and warehousing |
| 51 Information and cultural industries | Other service industries |
| 52 Finance and insurance | Other service industries |
| 53 Real estate and rental and leasing | Other service industries |
| 54 Professional, scientific and technical services | Other service industries |
| 55 Management of companies and enterprises | Other service industries |
| 56 Administrative and support, waste management and remediation services | Other service industries |
| 61 Educational services | Educational services  Excludes: 6116 - other schools and instruction |
| 62 Health care and social assistance | Health care and social assistance |
| 71 Arts, entertainment and recreation | Other service industries  Excludes: 7111 - performing arts companies, 7112 - spectator sports, 7113 - promoters (presenters) of performing arts, sports and similar events, 7114 - agents and managers for artists, athletes, entertainers and other public, 7115 - independent artists, writers and performers, 7131 - amusement parks and arcades 7132 - gambling industries, 7139 - other amusement and recreation industries |
| 72 Accommodation and food services | Accommodation and food services |
| 81 Other services (except public administration) | Other service industries  Excludes: 8131 - religious organizations |
| 91 Public administration | Public administration |

**Supplementary Appendix 2.** Flow diagram of study cohort

COVID-19 Cases reported between April 1, 2020 - March 31, 2021 (N=351,398)

- (N=351, 398)

Excluded cases <15 or ≥70 years of age (N=68,859)

COVID-19 cases in individuals aged 15-69 years (N=282,539)

Non workplace outbreak associated cases (N=247,371)

Workplace outbreak associated cases (N=35,168)

Case among residents of congregate care (N=4,026)

Non-outbreak related cases (N=234,169)

Outbreak-associated non-workplace cases (N=9,176)

**Supplementary Appendix 3: Socio-demographic characteristics of case and hospitalizations workplace outbreak-associated cases, non-outbreak related cases, residents in congregate care cases and outbreak-associated non-workplace cases among 15-69 year olds in Ontario, Canada**

|  | **Cases** | | | | **Hospitalizations** | | | |
| --- | --- | --- | --- | --- | --- | --- | --- | --- |
|  | **Workplace outbreak-associated** | **Non-outbreak related** | **Residents in congregate care** | **Outbreak-associated non-workplace** | **Workplace outbreak-associated** | **Non-outbreak related** | **Residents in congregate care** | **Outbreak-associated non-workplace** |
| **Total (Ontario)** | 35,168 | 234,169 | 4,026 | 9,176 | 557 | 6,122 | 456 | 798 |
| **Time** |  |  |  |  |  |  |  |  |
| Period 1 (April 1st - Aug 31st 2020) | 6,648 | 20,488 | 1,214 | 1,019 | 187 | 1,457 | 236 | 188 |
| Period 2 (Sept 1st - Dec 31st 2020) | 12,995 | 100,860 | 931 | 3,334 | 130 | 1,882 | 97 | 226 |
| Period 3 (Jan 1st - March 31st 2021) | 15,525 | 112,821 | 1,881 | 4,823 | 240 | 2,783 | 123 | 384 |
| **Gender** |  |  |  |  |  |  |  |  |
| Female | 19,534 | 113,355 | 1,268 | 4,584 | 243 | 2,564 | 188 | 298 |
| Male | 15,397 | 119,668 | 2,721 | 4,493 | 311 | 3,540 | 268 | 497 |
| Other* | 237 | 1,146 | 37 | 99 | 3 | 18 | 0 | 3 |
| **Age (years)** |  |  |  |  |  |  |  |  |
| 15-24 | 4,245 | 50,264 | 232 | 2,085 | 9 | 217 | 5 | 17 |
| 25-34 | 8,400 | 55,643 | 590 | 1,769 | 48 | 555 | 19 | 55 |
| 35-44 | 7,544 | 41,394 | 537 | 1,449 | 72 | 738 | 26 | 75 |
| 45-54 | 8,089 | 40,692 | 569 | 1,626 | 183 | 1,352 | 59 | 156 |
| 55-64 | 6,023 | 35,859 | 1,150 | 1,670 | 206 | 2,141 | 172 | 280 |
| 65-69 | 867 | 10,317 | 948 | 577 | 39 | 1,119 | 175 | 215 |
| **Material Deprivation Quintile**** |  |  |  |  |  |  |  |  |
| 1 - low | 4,419 | 34,990 | 8 | 1,360 | 57 | 731 | 0 | 100 |
| 2 | 5,800 | 37,155 | 6 | 1,529 | 110 | 846 | 1 | 118 |
| 3 | 6,765 | 43,960 | 4 | 1,405 | 115 | 1,007 | 1 | 121 |
| 4 | 7,424 | 48,008 | 6 | 1,620 | 115 | 1,187 | 0 | 158 |
| 5 - high | 8,607 | 56,411 | 9 | 2,224 | 145 | 2,011 | 1 | 223 |
| Missing | 2,153 | 13,250 | 2 | 1,032 | 15 | 291 | 0 | 78 |
| **Diversity Quintile**** |  |  |  |  |  |  |  |  |
| 1 - low | 2,700 | 11,040 | 9 | 844 | 36 | 327 | 0 | 64 |
| 2 | 4,045 | 16,031 | 3 | 1,127 | 74 | 421 | 0 | 111 |
| 3 | 4,445 | 25,135 | 4 | 1,433 | 75 | 655 | 1 | 130 |
| 4 | 6,550 | 46,199 | 7 | 1,822 | 116 | 1,126 | 0 | 173 |
| 5 - high | 15,275 | 122,119 | 10 | 2,912 | 241 | 3,235 | 2 | 242 |
| Missing | 2,153 | 13,250 | 2 | 1,032 | 15 | 291 | 0 | 78 |
| **Public Health Unit** |  |  |  |  |  |  |  |  |
| Algoma District | 16 | 142 | 2 | 35 | 1 | 1 | 0 | 0 |
| Brant County | 187 | 1,518 | 19 | 72 | 1 | 24 | 1 | 5 |
| Chatham-Kent | 295 | 968 | 3 | 50 | 2 | 20 | 0 | 2 |
| City Of Hamilton | 1,344 | 7,998 | 208 | 527 | 23 | 213 | 33 | 82 |
| City Of Ottawa | 1,703 | 10,427 | 447 | 438 | 37 | 291 | 50 | 42 |
| Durham Region | 1,592 | 9,381 | 145 | 203 | 31 | 256 | 10 | 18 |
| Eastern Ontario | 275 | 1,962 | 33 | 104 | 4 | 66 | 3 | 11 |
| Grey Bruce | 81 | 535 | 5 | 17 | 4 | 10 | 0 | 1 |
| Haldimand-Norfolk | 448 | 797 | 14 | 35 | 11 | 24 | 0 | 0 |
| Haliburton, Kawartha, Pine Ridge | 126 | 630 | 22 | 46 | 3 | 17 | 2 | 0 |
| Halton Region | 909 | 6,882 | 292 | 207 | 15 | 130 | 12 | 10 |
| Hastings & Prince Edward Counties | 82 | 327 | 3 | 15 | 3 | 8 | 0 | 0 |
| Huron Perth | 186 | 780 | 33 | 24 | 1 | 14 | 0 | 1 |
| Kingston, Frontenac, Lennox & Addington | 121 | 571 | 1 | 46 | 3 | 6 | 0 | 1 |
| Lambton County | 235 | 1,819 | 56 | 91 | 2 | 25 | 1 | 2 |
| Leeds, Grenville And Lanark District | 193 | 547 | 16 | 71 | 7 | 20 | 0 | 3 |
| Middlesex-London | 896 | 4,456 | 97 | 425 | 7 | 127 | 11 | 36 |
| Niagara Region | 1,748 | 4,920 | 142 | 517 | 31 | 113 | 15 | 27 |
| North Bay Parry Sound District | 16 | 161 | 3 | 57 | 0 | 14 | 0 | 4 |
| Northwestern | 26 | 295 | 3 | 208 | 1 | 15 | 0 | 10 |
| Southwestern | 498 | 1,582 | 33 | 40 | 6 | 39 | 4 | 5 |
| Peel Region | 7,272 | 50,803 | 248 | 708 | 74 | 860 | 32 | 41 |
| Peterborough County-City | 61 | 558 | 7 | 82 | 0 | 15 | 1 | 1 |
| Porcupine | 43 | 181 | 14 | 11 | 1 | 9 | 3 | 1 |
| Renfrew County And District | 85 | 224 | 1 | 8 | 0 | 5 | 0 | 0 |
| Simcoe Muskoka District | 1,123 | 4,874 | 66 | 210 | 35 | 145 | 3 | 23 |
| Sudbury And District | 203 | 728 | 6 | 131 | 3 | 25 | 2 | 7 |
| Thunder Bay District | 216 | 1,415 | 159 | 455 | 3 | 71 | 1 | 23 |
| Timiskaming | 22 | 70 | 2 | 8 | 0 | 4 | 2 | 2 |
| Toronto | 7,933 | 76,322 | 1,360 | 2,334 | 158 | 2,520 | 192 | 301 |
| Waterloo Region | 1,445 | 7,576 | 112 | 533 | 16 | 178 | 22 | 37 |
| Wellington-Dufferin-Guelph | 852 | 3,107 | 44 | 226 | 11 | 78 | 5 | 11 |
| Windsor-Essex County | 2,724 | 7,851 | 295 | 335 | 25 | 227 | 29 | 26 |
| York Region | 2,212 | 23,762 | 135 | 907 | 38 | 552 | 21 | 65 |

*Includes individuals for which gender was not reported or missing, as well as individuals reporting transgender or non-binary gender

**Quintile 5 represents the highest quintile of deprivation or diversity. The material deprivation measure combines information on income, quality of housing, educational attainment and family structure characteristics to assess the ability of individuals and communities to access and attain basic material needs. The ethnic concentration dimension is based on the proportion of non-white and non-Indigenous residents and/or the proportion of immigrants that arrived in Canada within the past five years.

**Supplementary Appendix 4: Cumulative case rate (per 100,000 workers) of COVID-19 among Ontario workers aged 15-69 by time and industry**

Rate (per 100,000 workers)

|  | LFS Cases per 100,000 workers (home=0) |
| --- | --- |
| Period 1 (April 1st - Aug 31st 2020) Agriculture, forestry, fishing and hunting | 2597.17 |
| Period 1 (April 1st - Aug 31st 2020) Manufacturing - Food | 477.471 |
| Period 1 (April 1st - Aug 31st 2020) Health care and social assistance | 761.663 |
| Period 1 (April 1st - Aug 31st 2020) Transportation and warehousing | 71.1728 |
| Period 1 (April 1st - Aug 31st 2020) Educational services | 102.045 |
| Period 1 (April 1st - Aug 31st 2020) Manufacturing - Other | 79.8166 |
| Period 1 (April 1st - Aug 31st 2020) Public administration | 22.2847 |
| Period 1 (April 1st - Aug 31st 2020) Accommodation and food services | 25.3165 |
| Period 1 (April 1st - Aug 31st 2020) Wholesale trade | 4.77983 |
| Period 1 (April 1st - Aug 31st 2020) Construction | 12.413 |
| Period 1 (April 1st - Aug 31st 2020) Retail trade | 7.66372 |
| Period 1 (April 1st - Aug 31st 2020) Mining and Utilities | 49.5946 |
| Period 1 (April 1st - Aug 31st 2020) Other Service Industries | 11.2875 |
| Period 2 (Sept 1st - Dec 31st 2020) Agriculture, forestry, fishing and hunting | 968.293 |
| Period 2 (Sept 1st - Dec 31st 2020) Manufacturing - Food | 959.921 |
| Period 2 (Sept 1st - Dec 31st 2020) Health care and social assistance | 876.451 |
| Period 2 (Sept 1st - Dec 31st 2020) Transportation and warehousing | 386.45 |
| Period 2 (Sept 1st - Dec 31st 2020) Educational services | 295.634 |
| Period 2 (Sept 1st - Dec 31st 2020) Manufacturing - Other | 301.432 |
| Period 2 (Sept 1st - Dec 31st 2020) Public administration | 100.796 |
| Period 2 (Sept 1st - Dec 31st 2020) Accommodation and food services | 165.805 |
| Period 2 (Sept 1st - Dec 31st 2020) Wholesale trade | 171.456 |
| Period 2 (Sept 1st - Dec 31st 2020) Construction | 45.1804 |
| Period 2 (Sept 1st - Dec 31st 2020) Retail trade | 79.8943 |
| Period 2 (Sept 1st - Dec 31st 2020) Mining and Utilities | 33.4746 |
| Period 2 (Sept 1st - Dec 31st 2020) Other Service Industries | 61.659 |
| Period 3 (Jan 1st - March 31st 2021) Agriculture, forestry, fishing and hunting | 1922.43 |
| Period 3 (Jan 1st - March 31st 2021) Manufacturing - Food | 1045.98 |
| Period 3 (Jan 1st - March 31st 2021) Health care and social assistance | 867.775 |
| Period 3 (Jan 1st - March 31st 2021) Transportation and warehousing | 619.791 |
| Period 3 (Jan 1st - March 31st 2021) Educational services | 446.493 |
| Period 3 (Jan 1st - March 31st 2021) Manufacturing - Other | 485.301 |
| Period 3 (Jan 1st - March 31st 2021) Public administration | 311.514 |
| Period 3 (Jan 1st - March 31st 2021) Accommodation and food services | 156.943 |
| Period 3 (Jan 1st - March 31st 2021) Wholesale trade | 175.573 |
| Period 3 (Jan 1st - March 31st 2021) Construction | 149.065 |
| Period 3 (Jan 1st - March 31st 2021) Retail trade | 114.063 |
| Period 3 (Jan 1st - March 31st 2021) Mining and Utilities | 139.368 |
| Period 3 (Jan 1st - March 31st 2021) Other Service Industries | 109.213 |

**Supplementary Appendix 5: Supporting data for Figure 1 and Supplementary Appendix 4**

|  |  | **Total Number of Hours** | | **Average Number of Workers** | |
| --- | --- | --- | --- | --- | --- |
| **Time period & Industry** | **Cases** | **Total Number of Hours Worked (majority hours outside the home)** | **Case rate per 100,000,000 hours worked**  **(majority hours outside the home)** | **Average number of workers (majority hours outside the home)** | **Case rate per 100,000 workers (majority hours outside the home)** |
| **Period 1 (April 1st - Aug 31st 2020)** |  |  |  |  |  |
| Agriculture, forestry, fishing and hunting | 1,339 | 48,153,959 | 2,781* | 51,556 | 2,597 |
| Manufacturing - Food | 474 | 84,207,299 | 563 | 99,273 | 477 |
| Health care and social assistance | 4,050 | 390,015,763 | 1,038 | 531,731 | 762 |
| Transportation and warehousing | 164 | 187,868,797 | 87 | 230,425 | 71 |
| Educational services | 45 | 33,429,083 | 135 | 44,098 | 102 |
| Manufacturing - Other | 313 | 341,111,765 | 92 | 392,149 | 80 |
| Public administration | 32 | 115,840,750 | 28 | 143,596 | 22 |
| Accommodation and food services | 49 | 123,507,904 | 40 | 193,549 | 25 |
| Wholesale trade | 6 | 104,561,653 | 6 | 125,528 | 5 |
| Construction | 43 | 293,157,284 | 15 | 346,412 | 12 |
| Retail trade | 42 | 377,679,497 | 11 | 548,037 | 8 |
| Mining and Utilities | 21 | 39,578,589 | 53 | 42,343 | 50 |
| Other Service Industries | 70 | 471,204,619 | 15 | 620,155 | 11 |
| **Period 2 (Sept 1st - Dec 31st 2020)** |  |  |  |  |  |
| Agriculture, forestry, fishing and hunting | 532 | 39,126,580 | 1,360* | 54,942 | 968 |
| Manufacturing - Food | 861 | 58,594,099 | 1,469 | 89,695 | 960 |
| Health care and social assistance | 5,862 | 394,230,318 | 1,487 | 668,834 | 876 |
| Transportation and warehousing | 1,153 | 190,001,785 | 607 | 298,357 | 386 |
| Educational services | 923 | 194,025,987 | 476 | 312,211 | 296 |
| Manufacturing - Other | 1,577 | 364,638,983 | 432 | 523,169 | 301 |
| Public administration | 130 | 81,801,155 | 159 | 128,973 | 101 |
| Accommodation and food services | 528 | 145,406,278 | 363 | 318,445 | 166 |
| Wholesale trade | 233 | 89,878,386 | 259 | 135,895 | 171 |
| Construction | 192 | 290,645,426 | 66 | 424,963 | 45 |
| Retail trade | 528 | 339,150,494 | 156 | 660,874 | 80 |
| Mining and Utilities | 19 | 41,960,750 | 45 | 56,759 | 33 |
| Other Service Industries | 457 | 450,104,813 | 102 | 741,174 | 62 |
| **Period 3 (Jan 1st - March 31st 2021)** |  |  |  |  |  |
| Agriculture, forestry, fishing and hunting | 705 | 18,796,210 | 3,751* | 36,672 | 1,922 |
| Manufacturing - Food | 991 | 47,740,439 | 2,076 | 94,744 | 1,046 |
| Health care and social assistance | 5,471 | 280,224,605 | 1,952 | 630,463 | 868 |
| Transportation and warehousing | 1,739 | 135,208,416 | 1,286 | 280,578 | 620 |
| Educational services | 1,138 | 114,756,384 | 992 | 254,875 | 446 |
| Manufacturing - Other | 2,450 | 257,680,749 | 951 | 504,841 | 485 |
| Public administration | 376 | 61,268,514 | 614 | 120,701 | 312 |
| Accommodation and food services | 391 | 89,073,950 | 439 | 249,135 | 157 |
| Wholesale trade | 221 | 64,267,322 | 344 | 125,874 | 176 |
| Construction | 562 | 187,336,542 | 300 | 377,017 | 149 |
| Retail trade | 718 | 245,766,795 | 292 | 629,478 | 114 |
| Mining and Utilities | 75 | 30,452,374 | 246 | 53,814 | 139 |
| Other Service Industries | 688 | 290,377,823 | 237 | 629,961 | 109 |

* Two sensitivity analyses related to the Agriculture, forestry, fishing and hunting industry were performed. 1) Accounting for temporary foreign workers (TFWs) decreased the COVID-19 incidence rates in the agriculture industry from 2,781 to 1,858 (period 1), 1360 to 927 (period 2), and from 3,751 to 2,208 (period 3) cases per 100,000,000 hours in those who worked mostly outside the home. 2) When we reclassified the hours of those self-employed (with employees) on farms to working outside the home (i.e., to ensure their exposure to others was enumerated), the incidence decreased across all time periods (to 2,275, 1,175, and 3,118 per 100,000,000 hours in periods 1-3, respectively).
